# Widening racial and ethnic disparities in *Helicobacter pylori* infection in the United States: a Bayesian meta-analytic catalytic model of age-specific forces of infection

**DOI:** 10.64898/2026.11.16.26367144

**Authors:** Tigist F. Menkir, Jorge A. Roa-Contreras, Yadira Peralta, Chelsea S. Taylor, Mercedes V. McMahon, Jennifer M. Yeh, M. Constanza Camargo, Robert J. Huang, Uri Ladabaum, Fernando Alarid-Escudero

## Abstract

**Background:** *Helicobacter pylori* is the main cause of non-cardia gastric cancer. In the U.S. *H. pylori* seroprevalence has declined. However, whether all subgroups have benefited equally from this change is unclear. Further, prevalence alone cannot indicate when infection risk is most acute and thus when to target prevention efforts like screening.

**Methods:** We synthesized age-stratified *H. pylori* seroprevalence data from 36 U.S. studies (N=56,845; 1965-2014), identified via literature search through September 2022. We fitted Bayesian hierarchical catalytic epidemic models to estimate, by race/ethnicity (for each of five race/ethnicity groups) and birth cohort (pre-1960 vs. 1960+, or continuous), the maximum proportion infected, age-specific force of infection (FOI), and the age at half-maximum seroprevalence.

**Findings:** Disparities in maximum prevalence worsened in the 1960+ birth cohort, with the absolute Hispanic-Non-Hispanic White (NHW) gap widening from 33.2 to 51.4 percentage points. FOIs peaked at the youngest ages for the Non-Hispanic American Indian/Alaska Native (AI/AN) and Non-Hispanic Black (NHB) populations. Age patterns varied independently of burden; Hispanic and AI/AN populations reached similar peak prevalence (83.0% vs 76.1%) for the 1960+ birth cohort, yet the age at half-maximum seroprevalence was 19.3 years (95%CrI: 14.9, 23.5) vs 3.20 years (95%CrI: 1.99, 4.63). Continuous birth-cohort analyses supported these findings.

**Interpretation:** National declines in *H. pylori* mask widening racial and ethnic disparities. In AI/AN and NHB populations, seroprevalence rises steeply in early childhood, well before adulthood-targeted screen-and-treat programs would intervene. Moreover, similar peak prevalence can mask different age patterns of infection, and should be accounted for when guiding prevention strategies.

**Funding:** National Cancer Institute U01CA265729 and U01CA319219

**Research in Context:** *Evidence before this study:* We used studies reporting *Helicobacter pylori* seroprevalence in U.S. populations from a recent systematic scoping review covering publications through 2022. Reported U.S. seroprevalence has declined over successive birth cohorts, and prevalence is consistently higher among Hispanic, Non-Hispanic Black, American Indian and Alaska Native, and Asian and Pacific Islander populations than among non-Hispanic White populations. Recent estimates mainly derive from limited populations, such as veterans or single integrated health systems. American Indian, Alaska Native, Asian, and Pacific Islander participants are also often grouped into a single “other” category. Catalytic epidemic models have been used to estimate age-specific force of infection for *H. pylori* in Mexico, Japan, and in a recent analysis of two NHANES cycles. However, none of these studies have estimated forces of infection separately for all five Surveillance, Epidemiology, and End Results program (SEER) racial and ethnic groups in the U.S. or across more than two survey periods. There has also been a lack of quantitative evidence for differences in the approximate age distribution of infection risk across race/ethnic groups and birth cohorts and how racial disparities in infection burden have changed over time.

*Added value of the study:* We applied modified catalytic epidemic models to estimate age-specific prevalence and rates of infection for each of the five race/ethnic SEER categories and by birth cohort. To additionally probe differences in when infection is typically concentrated by race/ethnicity and birth cohort group, we computed the age at which each group reaches half its maximum seroprevalence. Our approach pools data across distinct study sources and populations, accommodates study-specific heterogeneity and variation between race/ethnicity groups, and improves estimates in data-limited groups. Importantly, we improve the robustness of our key findings by applying a range of model extensions that accommodate distinct latent assumptions in catalytic epidemic models, namely the fundamental nature of the relationship between forces of infection and age and birth cohort effects. Our results reaffirm notable absolute and relative disparities in predicted prevalence and the risk of infection among race/ethnic minority groups, which have been intensified in more recent birth cohorts. We also demonstrate that infection trajectories maintain relatively distinct (early-age or later-onset) age trends across race/ethnic groups and within the same subgroup across birth cohorts.

*Implications of all the available evidence:* There is an ever-increasing need for evaluating expanded screen-and-treat programs for *H. pylori*, as population-level declines in prevalence have concealed notable *worsening* disparities in burden across race/ethnic groups. Screening strategies should be tailored to population-specific age-specific patterns in infection risk, with testing prioritized in early life where seroprevalence rises steeply in childhood, as in AI/AN and Non-Hispanic Black populations, and at first clinical contact for immigrants from high-incidence regions, whose infections likely predate arrival.

## Introduction

*Helicobacter pylori* (*H. pylori*), a bacterium known to trigger the inflammation-cancer pathway known as Correa’s cascade^1^, is a principal driver of gastric cancer globally.^2,3^ *H. pylori* is responsible for 85% of all non-cardia gastric cancer cases^2^ and is expected to contribute to 76% of gastric cancer incidence among younger birth cohorts, following an increasing trend in *H. pylori*-attributable cancers.^3^ Transmission, which occurs primarily through person-to-person interactions, is largely concentrated among socially disadvantaged populations.^4,5^ In the U.S., many of these communities also face an overlapping disproportionate burden of gastric cancer.^6^ Due to challenges related to access to care and information,^7^ *H. pylori* often remains underdetected and gastric cancer undiagnosed until later, more severe stages in these populations.^8^

*H. pylori* incidence typically follows an age-based trend, with the risk of infection most acute among youth. Within the U.S., the extent to which forces of infection and prevalence trend with age varies considerably across race/ethnicity groups, birth cohorts, and geographic locations.^9–11^ Burden is highest among race/ethnic minority populations and tends to be lower for younger birth cohorts, although these declines are not consistent across groups.^12^ As a consequence, in addition to facing an elevated risk of gastric cancer overall^6^, racial/ethnic minority populations, such as Hispanic Americans and Non-Hispanic Asian Americans, are reporting increasing rates of early-onset gastric cancer.^13^ The 2024 American College of Gastroenterology (ACG) guidelines recommend *H. pylori* testing in defined clinical circumstances, such as peptic ulcer disease, or among household contacts, and also incorporate race/ethnicity and immigration from high-incidence regions as high-risk criteria.^10^ However, neither the ACG nor the 2015 American Society for Gastrointestinal Endoscopy^14^ guidelines specify the age at which testing should begin in these populations, highlighting a persistent gap in recommendations that cannot be addressed using prevalence data alone. In contrast, estimated forces of infection, which represent *rates* of infection, can be used to extract the approximate age window when individuals in a subgroup face elevated risks of infection and would benefit most from treatment.

Thus, in order to inform updated understandings of *H. pylori* disparities in infection dynamics and guide extended screen-and-treat policies on optimal ages to screen, we evaluated group-specific rates of infection across ages and birth cohorts in the U.S.^9^ To do so, we leveraged data from a scoping review of *H. pylori* serological data drawn from ethnically diverse populations across the country, pooling US-born and foreign-born individuals from 1965-2014. While recent serological studies have been largely restricted to specific populations, e.g. veterans^15^, locations, e.g. northern California^12^, or older time periods, e.g. the two NHANES cycles of data in 1988-1991 and 1999-2000, reporting data only among Non-Hispanic White, Hispanic, Non-Hispanic Black, and Other populations^16^, our estimates are based on a temporally, geographically, and demographically heterogeneous study base.^9^ We applied a series of flexible hierarchical catalytic epidemic models to estimate age-specific forces of infection (FOIs), maximum potential prevalence, ages at half-maximum seroprevalence, and disparities in predicted prevalence across race/ethnic groups and birth cohorts in the U.S. These measures illustrate both the relative magnitude and trajectory of annual infection rates by age and race/ethnic group and thus when to target group-specific screening policies.

Additionally, estimated forces of infection can help parameterize transmission dynamic models used to evaluate the potential impacts of infection-targeting interventions.

## Methods

### Data sources

We obtained *H. pylori* seroprevalence collected for an expanded scoping review and included the two cycles of available NHANES serological data.^9^ Further details can be found in the Supplementary Appendix.

### Data analysis

#### Measures

Our factors of interest were 1) each of the five Surveillance, Epidemiology, and End Results (SEER) program-defined race/ethnicity categories (Hispanic, Non-Hispanic White [NHW,] Non-Hispanic Black [NHB], Non-Hispanic Asian and Pacific Islander [API], and Non-Hispanic American Indian/Alaska Native [AI/AN])^17^ and 2) birth cohort, through a binary classification of pre vs >=1960 (selected to approximate major sanitation improvements in the U.S. while balancing data across subgroups) and through a continuous classification of birth cohort years.

Our primary outcomes included, for each age, race/ethnicity group, and birth cohort: predicted *H. pylori* seroprevalence and FOI. The FOI represents the annual rate of infection among susceptible individuals of a given age. In the context of an open population, the estimated FOI also includes seropositive individuals entering the population through immigration. We also computed predicted seroprevalence by age 5 and by age 90 (maximum prevalence) for each race/ethnic group and birth cohort. To evaluate disparities in maximum prevalence, we computed relative measures (prevalence ratios and the index of disparity^18^) and absolute measures (prevalence differences, in percentage points) by birth cohort category, using the NHW population as the referent subgroup. We report both because relative measures are sensitive to changes in the referent group, whose own maximum prevalence declined substantially between birth cohorts. Finally, for each race/ethnic group and birth cohort, we calculated the age at half-maximum seroprevalence, which equals the median age at acquisition in catalytic models when the population is closed.^19,20^ Because the study population includes both U.S.- and foreign-born individuals, these ages describe when seropositivity accumulates in the U.S. population, not where or when infection occurred.

#### Models

We fit Bayesian hierarchical models, which we term meta-analytic catalytic epidemic models, to the study-level subgroup-specific seropositivity estimates to estimate age-specific prevalence and FOI in each race/ethnic group by birth cohort. Extending on Muench’s^19^ standard catalytic epidemic model, which assumes a constant force of infection over time, and thus that the proportion infected in a given population follows an exponential relationship with age, our model accommodates differences in the FOI by birth cohort (Supplementary Appendix methods). We further accounted for between-study heterogeneity via a study-level random intercept and an overdispersion parameter θ, to absorb any residual variation (Supplementary Appendix methods). The hierarchical model structure we apply allows us to combine age-specific seropositivity data across the diverse data sources and race/ethnic groups captured, improving estimation in more data-limited groups.

We assume observed seroprevalence for each age group *i* and race/ethnic group *j* follows a beta-binomial distribution according to a mean cumulative probability of infection p_ij_ as a function of age (ie., F(a_ij_)), the number of individuals in each subgroup n_ij_, and an overdispersion parameter θ, as follows:

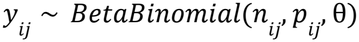

Equating F(a_ij_) with observed seroprevalence assumes no seroreversion (such that *prevalence* increases monotonically with age), no increased mortality risk among the infected, no migration into or out of the population, and that infection risk depends only on age and birth cohort and not otherwise on calendar time.

We note that the single-exponential cumulative form in our base model implies a force of infection that is maximal at birth and declines monotonically with age. This shape is a structural property of Muench’s formulation^19^ and implicitly assumes that infection risk peaks at birth. To test this assumption, we additionally fitted a generalized form which permits a non-monotone force of infection with a possible peak after birth. In our simplest formulation, we assumed that this shape is uniform across race/ethnicities. Under a more complex extension, we allowed shapes to vary across groups (See Supplementary Appendix).

Under our continuous birth cohort formulation, our base model assumes a linear relationship with birth cohort years. As an alternative, to borrow information from the recent past for sparse-data cohorts, we assumed birth cohort trends followed a random-walk structure, either with group-specific baselines and a shared trajectory or group-specific baselines *and* trajectories.

In our Supplementary Appendix methods, we detail all model specifications, priors, and equations for deriving force of infection and prevalence across these various formulations. We summarize each model through DIC and Pareto-smoothed importance sampling leave-one-out cross-validation (PSIS-LOO) (Supplementary Tables S1 and S2).

We implemented the “Just another Gibbs sampler” (JAGS) software in R to run Markov Chain Monte Carlo simulations and estimate posterior age- and race-specific forces of infection and prevalence by birth cohort category. For each chain, we ran an adaptation of 10,000 tuning iterations, with a burn-in of 500,000 iterations. We assessed convergence using the maximum univariate and multivariate potential scale reduction factor (PSRF) across parameters, effective sample sizes, and visual inspection of trace and density plots.

## Results

Model comparison metrics are reported in Supplementary Table S1. The selected binary cohort model was the base model with study random effects, and there was little evidence to favor the more complex formulations allowing for non-monotonic trends in FOI (Supplementary Tables S1 and S3). We selected the continuous cohort model with a random walk structure, group-specific intercepts, and population-wide trajectories (Supplementary Table S2). Both models showed evidence of robust parameter convergence (Supplementary Appendix Section D and Figures S1-S6). Predicted prevalence curves by birth cohort category generally aligned with observed prevalence estimates and their uncertainty ranges (Supplementary Figure S7). Notable non-overlap occurred among older ages in the pre-1960 birth cohort for the Hispanic population, where predictions systematically overestimated the observed data (Supplementary Figure S7).

Predicted prevalence by age 90 in the 1960+ birth cohort was highest in the Hispanic (83.0%, 95%CrI: 73.4%, 89.4%) and AI/AN (76.1%, 95%CrI: 57.5%, 90.3%) populations, followed by the NHB (63.7%, 95%CrI: 45.5%, 81.5%), API (45.9%, 95%CrI: 16.7%, 72.8%) and NHW (31.5%, 95%CrI: 15.4%, 43.9%) populations (Figure 1a). Estimates were least precise for the API population, for which only one study contributed to this cohort. Relative to the pre-1960 cohort, maximum prevalence declined in every group except the Hispanic population, where it increased slightly (3.22 percentage points, 95%CrI: −4.53, 10.3) (Figure 1a). Predicted prevalence by age 5 was highest in the AI/AN and NHB populations in the 1960+ birth cohort, consistent with earlier infection timing in these groups (Figure 1b). The birth-cohort-year analysis supported these findings, with the gap between prevalence at age 5 and age 90 more pronounced for earlier cohorts, consistent with later-age infection trends in the past (Figure 2).

**Figure 1.**
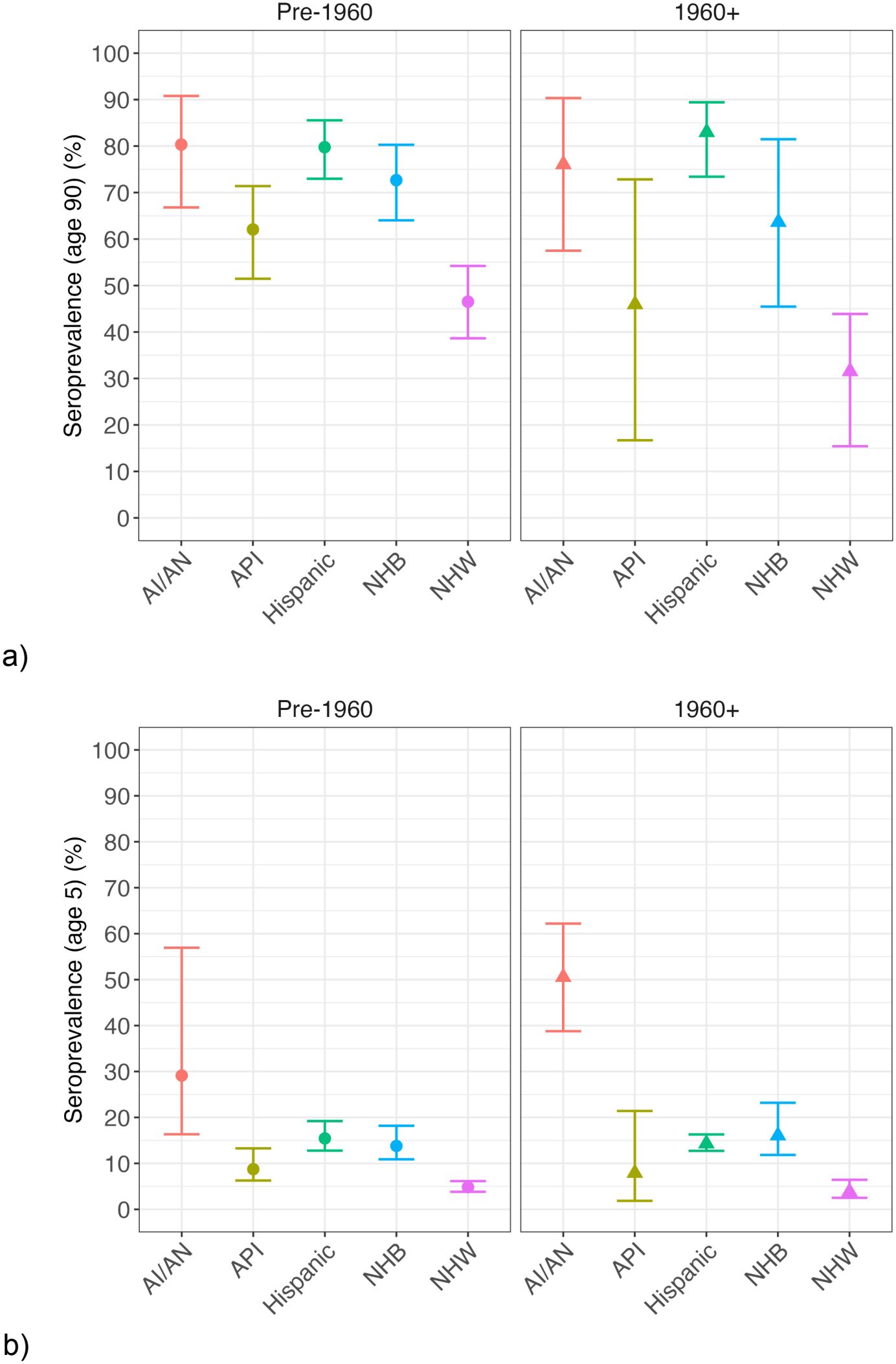
Model-predicted prevalence (%) by age 90 by race/ethnic group, stratified by birth cohort category (pre-1960 birth cohort (left, circles) and 1960+ birth cohort (right, triangles)) (a). Model-predicted prevalence (%) by age 5 by race/ethnic group, stratified by birth cohort category (pre-1960 birth cohort (left, circles) and 1960+ birth cohort (right, triangles)) (b). Points and error bars indicate posterior means and 95% confidence intervals. AI/AN: American Indian/Alaska Native, API: Asian and Pacific Islander, NHB: Non-Hispanic Black, NHW: Non-Hispanic White.

**Figure 2.**
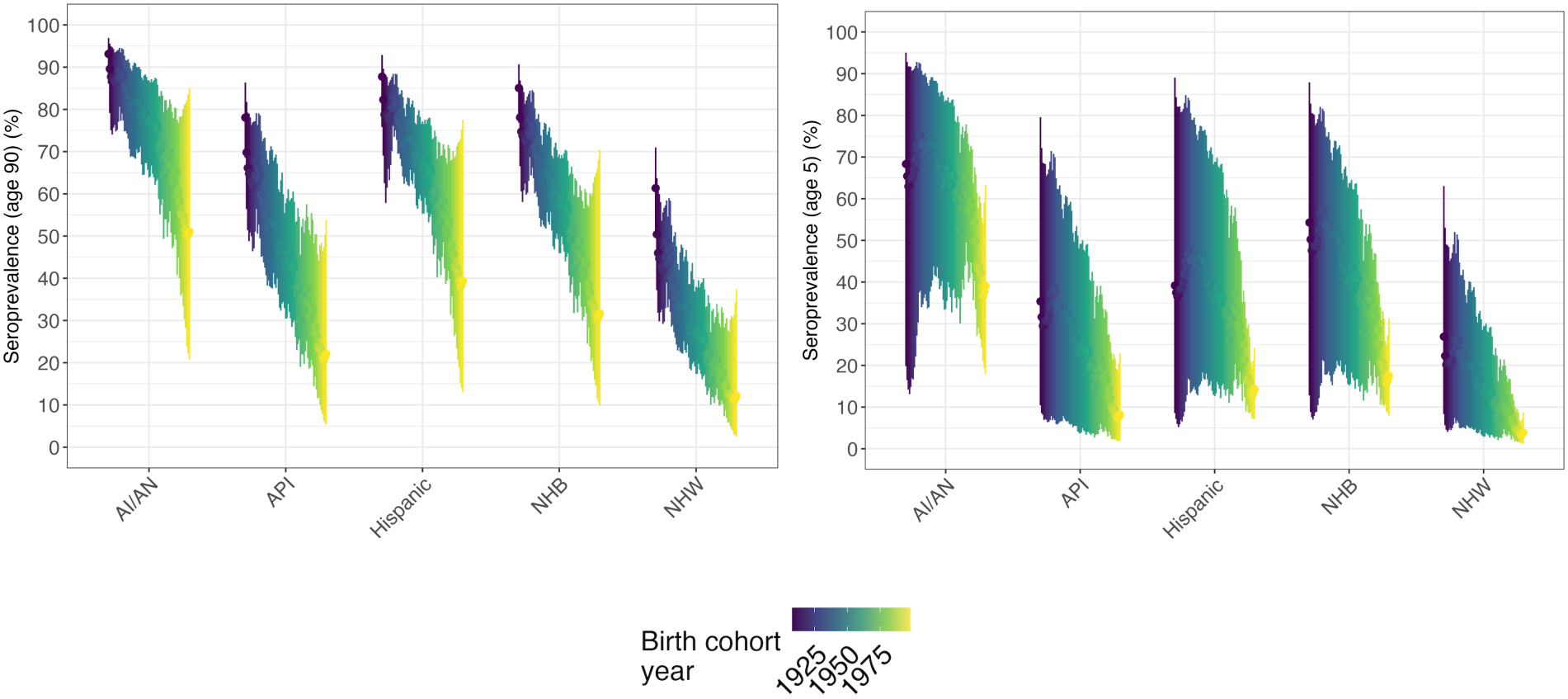
Model-predicted prevalence (%) by age 90 (left) and prevalence (%) by age 5 (right) by race/ethnicity group, across birth cohort years. Points and error bars indicate observed means and 95% confidence intervals. AI/AN: American Indian/Alaska Native, API: Asian and Pacific Islander, NHB: Non-Hispanic Black, NHW: Non-Hispanic White.

Disparity metrics reinforced substantial differences in maximum prevalence across race/ethnic groups, which worsened over time. For the pre-1960 birth cohort, prevalence ratios for the AIAN, Hispanic, NHB, and API populations compared to the NHW population were 1.74 (95%CrI: 1.39, 2.13), 1.72 (95%CrI: 1.54, 1.97), 1.57 (95%CrI: 1.41, 1.78), and 1.34 (95%CrI: 1.13, 1.58). These ratios increased for all groups in the 1960+ birth cohort to 2.59 (95%CrI: 1.58, 4.90), 2.83 (95%CrI: 1.92, 5.32), 2.13 (95%CrI: 1.38, 3.74), and 1.53 (95%CrI: 0.586, 2.96), respectively. The overall index of disparity also increased from 59.2 (95%CrI: 41.2, 83.0) to 129 (95%CrI: 60.3, 306). Absolute disparities widened alongside relative ones. The difference in maximum prevalence between the Hispanic and NHW populations increased from 33.2 percentage points (95% CrI: 28.1, 38.8) to 51.4 percentage points (95% CrI: 38.7, 68.4). The corresponding AI/AN-NHW difference increased from 33.8 percentage points (95% CrI: 19.3, 46.3) to 44.5 percentage points (95% CrI: 22.9, 65.2).

Estimated FOI curves further recapitulated these trends (Figure 3). The steepest declines in FOI from very elevated initial values at age = 1, were observed for the AI/AN and NHB groups, particularly for the 1960+ birth cohort, where FOI peaked at rates of 0.160/yr (95%CrI: 0.110/yr, 0.227/yr) and 0.0367/yr (95%CrI: 0.0255/yr, 0.0577/yr), respectively (Figure 3). The Hispanic, API, and NHW populations displayed peak rates of infection of 0.0311/yr (95%CrI: 0.0273/yr, 0.0361/yr), 0.0180/yr (95%CrI: 0.0038/yr, 0.0554/yr), and 0.0081/yr (95%CrI: 0.0051/yr, 0.0145/yr) in this birth cohort category (Figure 3). The 1960+ birth cohort of the Hispanic population reported a slightly increased risk at older ages compared to the pre-1960 birth cohort, while for all other populations, the opposite was true (Figure 3). The individual birth cohort year analysis revealed generally downshifted FOI curves across ages for the more recent birth cohorts for all groups, although this time trend was less consistent for the Hispanic, AI/AN, and NHB populations (Figure 4). Rapid early-age declines in rates of transmission were more pronounced for the younger birth cohorts, most acutely for the AI/AN and NHB populations, but less so for the Hispanic population.

**Figure 3.**
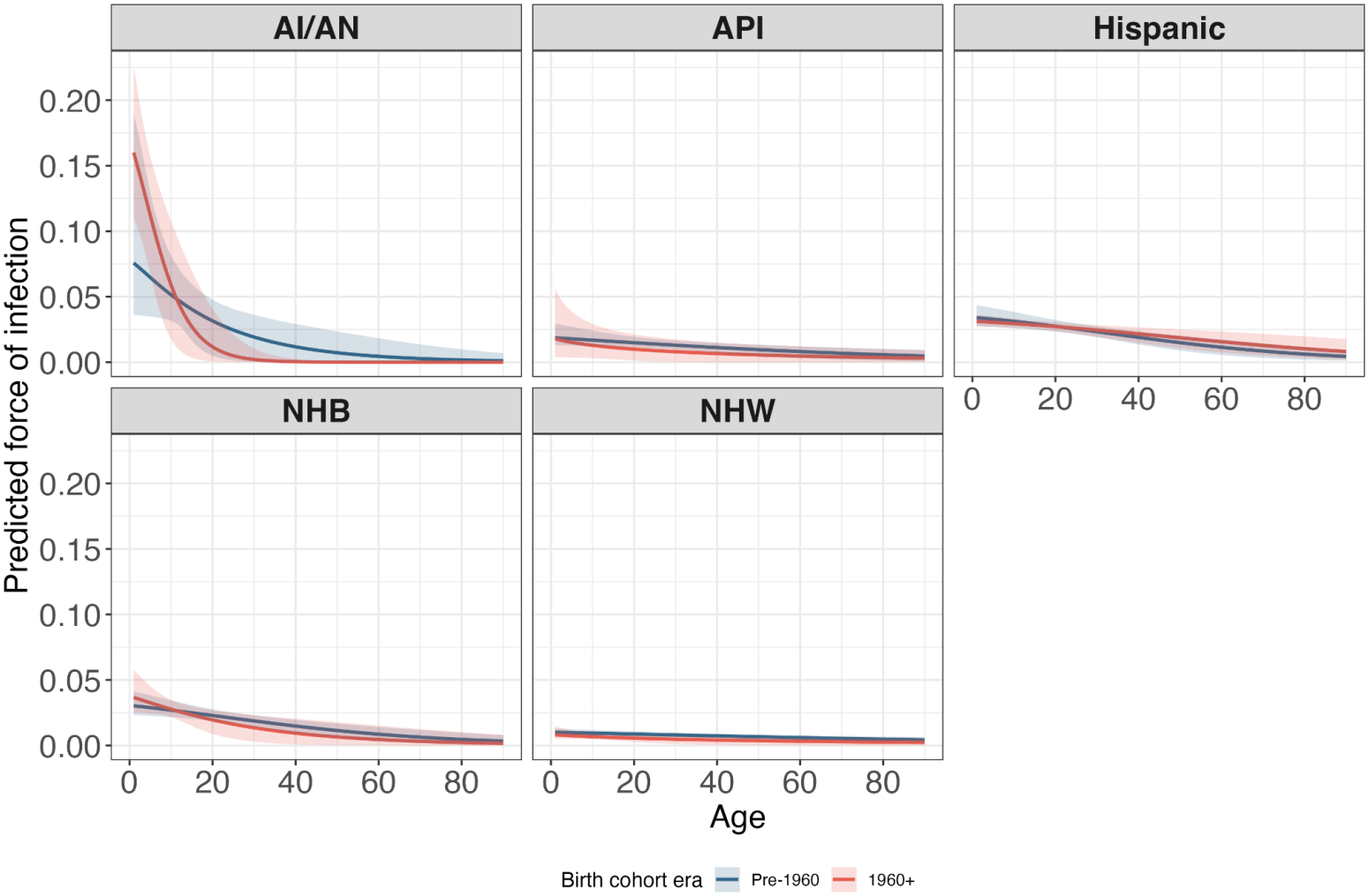
Model-predicted forces of infection by age, race/ethnicity group and birth cohort category (pre-1960 birth cohort (blue) and 1960+ birth cohort (red)). Curves indicate posterior means and shaded regions indicate 95% credible intervals. AI/AN: American Indian/Alaska Native, API: Asian and Pacific Islander, NHB: Non-Hispanic Black, NHW: Non-Hispanic White.

**Figure 4.**
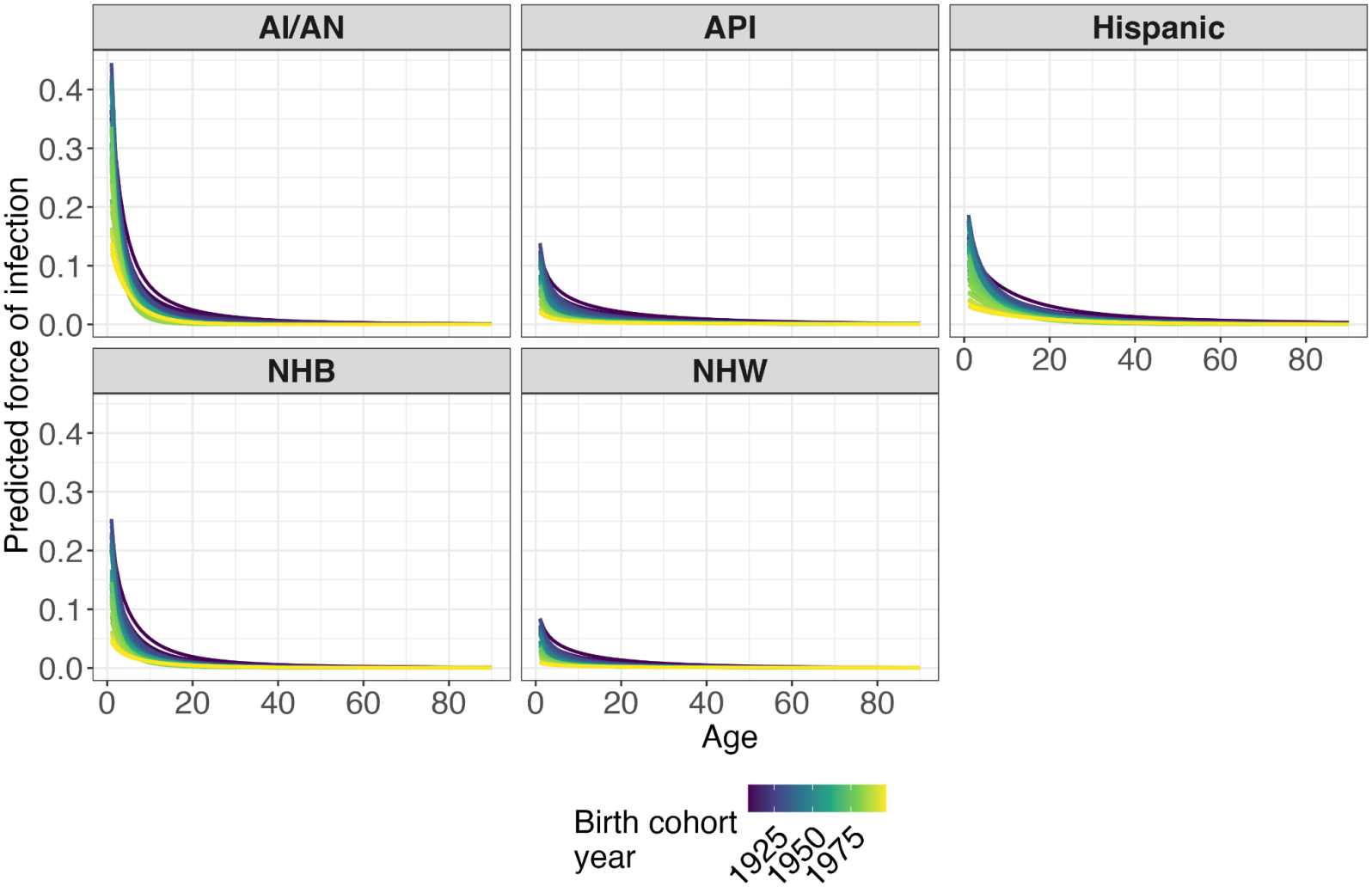
Model-predicted forces of infection by age, race/ethnic group and birth cohort years. Curves indicate posterior means. AI/AN: American Indian/Alaska Native, API: Asian and Pacific Islander, NHB: Non-Hispanic Black, NHW: Non-Hispanic White.

Finally, ages at half-maximum seroprevalence also differed across race/ethnic groups (Figure 5). Consistent with the generally increased risk of early infection among the AI/AN and NHB populations, particularly in the 1960+ birth cohort, we observed younger ages at half-maximum seroprevalence in these groups (3.20yrs, 95%CrI: 1.99yrs, 4.63yrs and 12.8yrs, 95%CrI: 5.69yrs, 22.1yrs) (Figure 5a). The opposite was true for the NHW, API, and Hispanic populations (42.5yrs, 95%CrI: 7.93yrs, 102yrs; 36.1yrs, 95% CrI: 3.78yrs, 124yrs; 19.3yrs, 95%CrI: 14.9yrs, 23.5yrs) (Figure 5a). Finally, consistent with the increased predicted forces of infection for older ages among the Hispanic population for the 1960+ birth cohort, the age at half-maximum seroprevalence increased in the 1960+ birth cohort from 16.8yrs, 95% CrI: 12.3yrs, 21.4yrs to 19.3yrs, 95%CrI: 14.9yrs, 23.5yrs (Figure 5). These ages did not uniformly decrease across birth-cohort years (Figure 5). For the Hispanic, NHB, and AI/AN populations, ages at half-maximum seroprevalence dropped sharply from the earliest to intermediate birth cohorts (Figure 5), but slightly increased in the most recent birth cohorts, particularly among Hispanic populations. Conversely, for the API and NHW populations, ages at half-maximum seroprevalence generally increased across birth cohort years, though this trend was punctuated by brief declines (Figure 5). This fluctuating pattern is reflected in the corresponding binary cohort results, where estimated ages at half-maximum seroprevalence in the 1960+ birth cohort showed considerable uncertainty in these groups.

**Figure 5.**
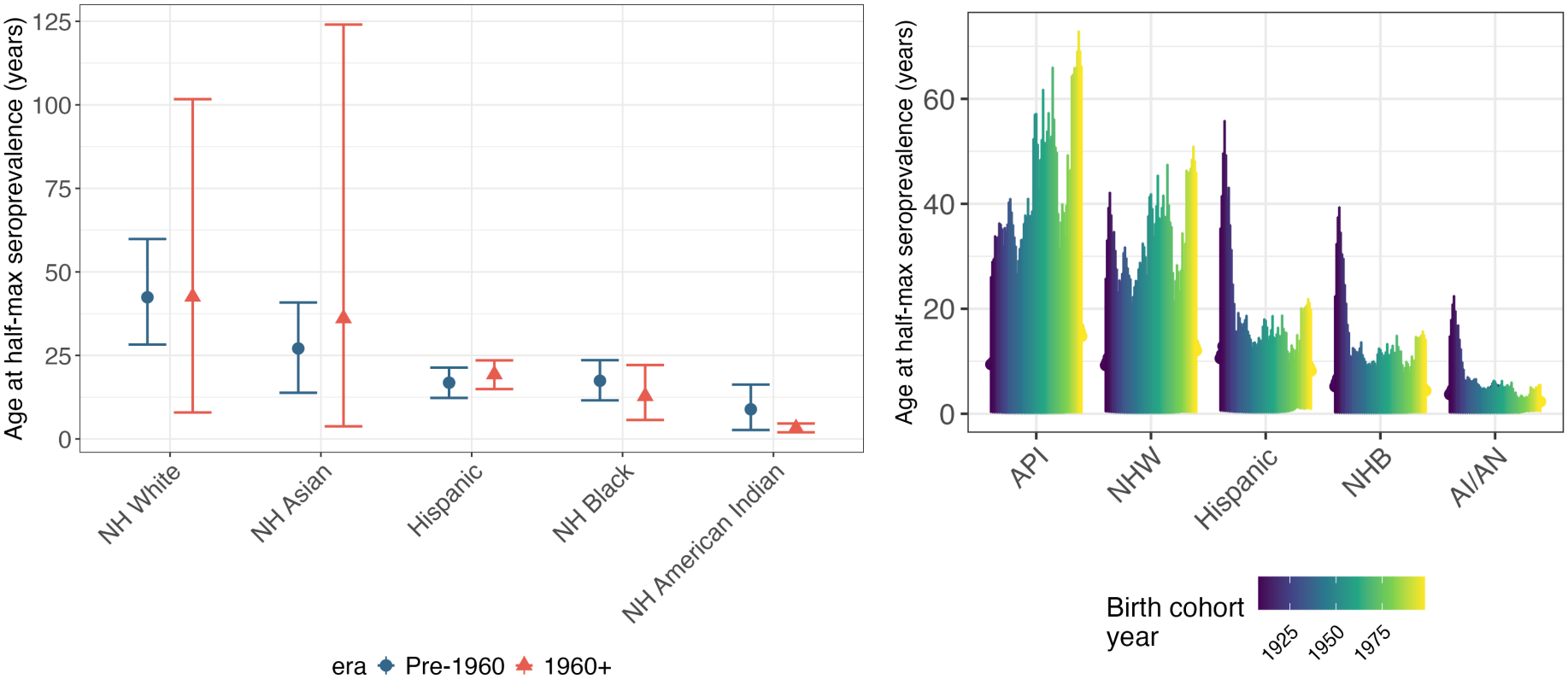
Model-predicted age at half-maximum seroprevalence (years) by race/ethnicity group and birth cohort category (pre-1960 birth cohort (pink, circles), 1960+ birth cohort (blue, triangles)) (left) or by birth cohort years (right). Points and error bars indicate posterior means and 95% confidence intervals. AI/AN: American Indian/Alaska Native, API: Asian and Pacific Islander, NHB: Non-Hispanic Black, NHW: Non-Hispanic White.

## Discussion

Our study provides the first age-specific estimates of *H. pylori* FOI for all five SEER race/ethnic groups covering five decades of birth cohorts. As such, we supply insights into differential patterns of the timing of infection across subgroups that cannot be deduced from a simple comparison of estimated prevalence. Two populations in our analysis carry similar lifetime *H. pylori* burden but reach half their lifetime seroprevalence more than fifteen years apart. Among individuals born in 1960 or later, predicted maximum prevalence was 83.0% and 76.1% in the Hispanic and AI/AN populations, yet the age at half-maximum seroprevalence was 19.3 years versus 3.2 years.

We also observed that *H. pylori* risk did not universally decrease among younger generations. For the younger birth cohort, predicted risk was increased among younger ages for the NHB and AI/AN populations and among older ages for the Hispanic population. For the NHB and AI/AN populations, FOI trends revealed accelerated early transmission, whereas for the Hispanic, API, and NHW populations, FOI declined more gradually with age.

The extent to which predicted prevalence by age 90 was reduced for younger birth cohorts also varied substantially across groups. For the Hispanic and AI/AN populations, maximum prevalence was generally stable across birth cohorts, indicating minimal headway in reducing *H. pylori* infection within these populations. In the Hispanic population, peak prevalence was higher in the younger birth cohorts, driven by the increased FOI predicted among older age groups. This trend may reflect migration rather than transmission within the U.S., as the Hispanic population includes a large proportion of recent immigrants^21^, among whom *H. pylori* is more prevalent.^16^

Finally, we found evidence for worsening disparities in all race/ethnic minority populations, with the index of disparity for maximum prevalence estimates more than doubling in the 1960+ birth cohort, with this widening concordant on the absolute scale.

Some of these differential trends provide insights into the appropriate time scale of action for targeting interventions in these subgroups. Among the AI/AN and NHB populations, where *H. pylori* risk appears to be acutely increased among very young ages, it may be important to ensure screen-and-treat programs are administered earlier on to reduce the likelihood of progression through gastric inflammation pathways by the time of first screen. This finding of the utility of potential early-age screening in these groups aligns with results from a microsimulation modeling study evaluating costs and benefits of targeted screening in a single birth cohort of NHW and NHB populations.^22^ For the Hispanic and API populations, older ages at half-maximum seroprevalence should not be interpreted as a reason to test later. Many individuals in these groups were likely infected in early childhood before immigrating, so testing foreign-born individuals from high-incidence regions at first clinical contact, as the ACG guidelines^10^ already allows, may matter more than any age threshold. Finally, acknowledging the critical role of within-household transmission in many of these communities, family-based treatment strategies, cited as effective tools for reducing individual and intrafamilial gastric cancer risk^23^, should also be considered..

Our analysis benefited from several key strengths. First, we utilized a Bayesian hierarchical framework previously developed and validated in other contexts.^16,24,25^ This approach bridges information about disease activity across subgroups to produce finer-resolution estimates of prevalence and forces of infection, increasing model stability in data-poor populations while maintaining distinct group-specific trends.^16,24,25^ Importantly, we collated data^9^ across disparate studies by explicitly accommodating differences in study design and *H. pylori* measurement. We tested additional model extensions reflecting varying assumptions about how forces of infection vary with age and potential group-specific birth cohort trends. We modeled birth cohorts both categorically and continuously, and our main findings held in both cases. By doing so, we extend earlier approaches outlined in Kamau et al.^20^, Kang et al.^26^, and Hay et al.^27^, namely non-monotonic, flexible trends in FOI, and additional modifications (e.g. explicit meta-analytic hierarchical model structure).

In the context of other recently published studies in the U.S., our results reported similar race/ethnicity disparities in predicted prevalence trends. A study conducted among veterans from 1999-2018 found higher seropositivity in NHB (40.2%), Hispanic (36.7%), and other populations (27.7%) than in NHWs (20.1%), with no improvements in these disparities over time.^15^ A study of Kaiser Permanente Northern California users from 2000-2019 also revealed race/ethnic disparities in serological positivity among Hispanics (56.4%), NHBs (49.0%), and APIs (42.5%) compared to NHWs (22.3%).^12^ Importantly, our age-specific estimates of seroprevalence in the API population (our most data-sparse group) broadly aligned with those reported in the study. Finally, a study conducted among an uninsured, predominantly Hispanic population (88.7%) in Southeastern Texas also demonstrated a high prevalence among Hispanics compared to NHWs: 65.9% versus 45.0% .^28^

Our study has several important limitations. First, the data did not distinguish US-born from foreign-born individuals, and our models assume a closed population. A person infected in early childhood abroad who immigrated as an adolescent or adult enters the data as seropositive at an older age. The estimated FOI therefore combines infection acquired in the U.S. with the arrival of already infected immigrants, and our data cannot separate the two. As previously noted, this may partly explain the older ages at half-maximum seroprevalence in the Hispanic and API populations, which have the largest foreign-born shares.^21^ For example, in Mexico, the main origin of Hispanic immigrants^21^, FOI is highest at birth, and the implied median age at infection is about 8 years.^24^ Older ages in the mostly US-born NHW population suggest migration is not the only explanation.

Additionally, our study sample for the API population was extremely sparse in younger birth cohorts, leading to imprecise estimates of prevalence and force of infection in these cohorts. It is thus imperative to invest more attention in increasing data representativeness in these birth cohorts for future studies, particularly given the high burden of non-cardia gastric cancer observed in the API population.^6^ Furthermore, racial/ethnic group categorization consolidates diverse ethnicities reporting very distinct risk profiles; for instance, seropositivity is consistently highest among Korean and Southeast Asian Americans.^12,29^ We also could not evaluate differences by sex, which has been clearly established as another important differentiating factor^30^, as this data was only available for a small sample of studies included in the scoping review.

Our catalytic epidemic model matches the fitted cumulative distribution of infection to the observed seroprevalence and therefore assumes no seroreversion. Seroreversion after spontaneous clearance or antibiotic eradication would cause observed seroprevalence to underestimate true cumulative infection with age, biasing the maximum prevalence downward and causing the observed curve to plateau earlier. This bias likely varies across birth cohorts with increasing exposure to eradication therapy. Finally, while we note that our 1960 birth cohort cutoff was partly arbitrary, the continuous cohort analysis supported the binary cohort findings.

Nevertheless, here we provide a rigorously tested, flexible framework yielding estimates of *H. pylori* prevalence and forces of infection across a range of race/ethnic groups, ages, and birth cohort categories in the U.S. Our findings suggest that claims of *population-wide* reductions in *H. pylori* infection risk over time in the U.S. hide important within-population race/ethnic disparities that are in fact worsening. We also show that age-based screening guidelines should account for subgroup variation in the timing of infection risk; if risk is concentrated in early childhood, earlier screening may be necessary to halt chronic gastric inflammation. Finally, we provide direct estimates of age-, race/ethnicity-, birth cohort-specific forces of infection that can be used to parameterize future modeling studies projecting the impacts of such targeted screening policies across at-risk populations.

## Data Availability

Input data and code necessary to run all models and produce numeric output and figures are available at https://github.com/goshgondar2018/HP_sero_hierarchical_model_US_expanded

https://github.com/goshgondar2018/HP_sero_hierarchical_model_US_expanded

## Contributors

TFM conceptualized the study, developed the methodology, wrote the software, did the formal analysis and visualization, and wrote the original draft. FA-E conceptualized the study, developed the methodology, supervised the work, acquired funding, and reviewed and edited the manuscript. YP developed the Bayesian catalytic methodology and reviewed and edited the manuscript. JAR-C curated the NHANES data, contributed software from the companion analysis, and reviewed and edited the manuscript. MVM and CST curated the scoping review data and reviewed and edited the manuscript. JMY and MCC contributed to the design and interpretation of the scoping review and critical review and editing of the manuscript. RJH and UL contributed to interpretation and critical review of the manuscript. TFM, JR, MVM, and CST accessed and verified the underlying data. All authors had full access to all study data and had final responsibility for the decision to submit for publication.

## Disclaimer

This research was supported in part by the Intramural Research Program of the National Institutes of Health (NIH). The contributions of the NIH authors are considered Works of the United States Government. The findings and conclusions presented in this paper are those of the authors and do not necessarily reflect the views of the NIH or the U.S. Department of Health and Human Services.

