## Supplementary Appendix for "Widening racial and ethnic disparities in *Helicobacter pylori* infection in the United States: a Bayesian meta-analytic catalytic model of age-specific forces of infection"

### A. Data

For the scoping review[1] we drew from, studies were selected based on whether they provided *H. pylori* serology age-annotated data for any of the five SEER race/ethnic groupings (Hispanic, Non-Hispanic White NHW, Non-Hispanic Black NHB, Non-Hispanic Asian and Pacific Islander API, and Non-Hispanic American Indian/Alaska Native AI/AN[2]). The total sample size, including two NHANES cycles of data, was n=56,845. NHANES data entered the models as unweighted counts of participants tested and testing positive in each age and race/ethnicity stratum, consistent with the other included studies, which report unweighted counts. We defined positive serological classifications using data from ELISA (the majority of studies) and Multiplex serology assays. We note that since the period during which the meta-analysis was conducted, two large seroprevalence studies in the U.S. have been published, namely Shah et al.[3] and Li et al.[4] We did not include these additional studies for the following reasons. Shah et al.[3] was conducted in a uniquely increased risk population (veterans[5]) and did not report seroprevalence across all five SEER race/ethnicity groups. Li et al.[4] was conducted in a specific geographic location and patient population (Kaiser Permanente Northern California users) and with its extremely large study sample size (544,000) would skew the results of our study towards this non-representative population. Nevertheless, we provide a detailed commentary of our results in the context of the findings presented in these and other studies in our Discussion section.

### B. Model specifications

<u>Underlying model equations</u>

$$F(a) = \alpha(1 - e^{-\gamma a}) \quad (1)$$
$$\lambda(a) = \frac{1}{1-F(a)}\frac{dF(a)}{da} \quad (2)$$

where $F(a)$ denotes the cumulative probability of infection by a given age a, $\lambda(a)$ denotes the force of infection at age a, $\alpha$ denotes the asymptote or maximum prevalence, and $\gamma$ denotes the rate of infection for each year of age a.

Given Equations 1 & 2, $\lambda(a)$ can be expressed as a function of $\alpha$ and $\gamma$ as follows :

$$\lambda(a) = \frac{\alpha\gamma e^{-\gamma a}}{1-\alpha(1-e^{-\gamma a})} \quad (3)$$

*Approach #1 (binary birth cohort of pre-1960 vs 1960+)*

1. <u>Base model</u>
   A. Model description

In the base model, the birth cohort binary is assumed to vary linearly with logit(alpha) and log(gamma), as follows. For this version, we do not account for study-level random effects and assume any extra-variation is only accounted for by the overdispersion parameter $\theta$.

$$logit(\alpha) = \alpha_0 + \alpha_1 BC.binary \quad (4)$$

$$\log(\gamma) = \gamma_0 + \gamma_1 BC.binary \quad (5)$$

where $BC.binary$=1 if birth cohort>=1960*

   B. Model priors

We placed a multivariate prior on the race-specific random effects $(\alpha_{kj}, \gamma_{kj})$ ( 6), weakly informative Cauchy priors on the $\alpha$ intercept and slope parameters (7 & 8) and $\gamma$ slope parameter (10). For the $\gamma$ intercept parameter, we imposed a Student's t prior informed by previous data on population-wide infection rates[6], to avoid unreasonably high estimated forces of infection (9). We assumed a Wishart prior for the precision (inverse covariance) matrix (11). Finally, we specified a relatively non-informative lognormal prior for the overdispersion parameter (12)

$$(\alpha_{kj}, \gamma_{kj}) \sim MVN((\alpha_{k,\mu}, \gamma_{k,\mu}), \Sigma_{\alpha\gamma}) \text{ where k denotes race/ethnic group } (6)$$

$$\alpha^{0}_{k,\mu} \sim Cauchy(0, 1) \quad (7)$$

$$\alpha^{1}_{k,\mu} \sim Cauchy(0, 1) \quad (8)$$

$$\gamma^{0}_{k,\mu} \sim t_{(3)}(-3.9, 1) \quad (9)$$

$$\gamma^{1}_{k,\mu} \sim Cauchy(0, 1) \quad (10)$$

$$\Sigma_{\alpha\gamma}^{-1} \sim W(I(4), 5) \quad (11)$$

$$\theta \sim Lognormal(3, 1) \quad (12)$$

2. <u>Base model with study random effects</u>
   A. Model description

In this model, we now include study-level random effects for the alpha parameter (reasoning that study-specific measurement and other differences are likely only to affect the overall level of prevalence, as opposed to the nature of infection trends) in addition to the overdispersion parameter $\theta$ to account for residual between-study variability, as follows:

$$logit(\alpha) = \alpha_0 + \alpha_1 BC.binary + \alpha_2 study.RE \quad (13)$$

B. Model priors

In addition to the priors specified in 6-12, we assumed the following distribution for study-level random effects:

$$u_s \sim Normal(0, \sigma_{study}^2) \ (14)$$

where we place a weakly informative inverse gamma prior on their variances:

$$\sigma_{study}^2 \sim IG(2, 0.5) \ (15)$$

3. <u>Model with common (global) kappa</u>

A. Model description

We expanded the base model to additionally accommodate non-monotonic changes in forces of infection over time, as follows:

$$F(a) = \alpha(1 - e^{-(\gamma a)^{\kappa}}) \quad (16)$$

$$\lambda(a) = \frac{\alpha\gamma\kappa(\gamma a)^{\kappa-1} e^{-(\gamma a)^{\kappa}}}{1-\alpha(1-e^{-(\gamma a)^{\kappa}})} \ (17)$$

The birth cohort binary is again assumed to vary linearly with logit(alpha) and log(gamma), as in model 1 (Equations 4 & 5)

B. Model priors

In addition to the priors specified in 6-12, we assumed the following lognormal distribution for kappa that is weakly informative and constrained to positive values:

$$\kappa \sim Lognormal(0, \tau = 1.5) \ (18)$$

4. <u>Model with common (global) kappa and study random effects</u>

A. Model description

This model extends on the global kappa model (model 3) by additionally including study random effects.

B. Model priors

All priors in 6-12, 14&15, and 18

5. <u>Model with group-specific kappas</u>

A. Model description

We expanded the kappa model to additionally accommodate variation in these non-monotonic changes in forces of infection across race/ethnic groups, as follows. Here, rather than explicitly set a different distribution to sample from for each group's kappa, we allow each group to maintain a distinct posterior value of kappa that varies from the population mean depending on the level of information they provide (sample sizes).

$$F(a) = \alpha(1 - e^{-(\gamma a)^{\kappa_k}})\ (19)$$

$$\lambda(a) = \frac{\alpha\gamma\kappa_k(\gamma a)^{\kappa_k - 1} e^{-(\gamma a)^{\kappa_k}}}{1-\alpha(1-e^{-(\gamma a)^{\kappa_k}})}\ (20)$$

The birth cohort binary is again assumed to vary linearly with logit(alpha) and log(gamma), as in model 1 (Equations 4 & 5)

B. Model priors

In addition to the priors specified in 6-12, we specify the prior for (now group-specific) $\kappa$ as follows:

$$log(\kappa) \sim N(\mu_{log(\kappa)}, \sigma_{log(\kappa)}^2)\ (21)$$

with weakly informative normal and gamma hyperpriors on $\mu$ and $\sigma^2$

$$\mu_{log(\kappa)} \sim N(0, 1)\ (22)$$

$$\sigma_{log(\kappa)}^2 \sim IG(2, 1)\ (23)$$

6. <u>Model with group-specific kappas and study random effects</u>

A. Model description

This model contains study-level random effects and group-specific kappas for differential trends in alpha and gamma by race/ethnicity.

B. Model priors

All priors in 6-12, 14&15, and 21-23

*Approach #2 (continuous birth cohort years)*

1. <u>Base model</u>

A. Model description

In the base model, continuous birth cohort years (which are centered based on the minimum across birth cohort years) is assumed to vary linearly with logit(alpha) and log(gamma), as follows. We also account for overdispersion and study random effects as before.

$$logit(\alpha) = \alpha_0 + \alpha_1 BC.continuous + \alpha_2 study.RE\ \ (24)$$

$$log(\gamma) = \gamma_0 + \gamma_1 BC.continuous\ \ (25)$$

where $BC.continuous$ denotes individual birth cohort years

B. Model priors

All priors in 6-12 and 14&15

2. <u>RW model with population-wide trajectories but group-specific intercepts</u>

A. Brief description

For the random walk model, observed birth cohort years are assumed to follow a random walk as they relate to both logit(alpha) and log(gamma), with common trajectories across groups but distinct starting points or intercepts at the baseline cohort. The model equations are the same as for the base model.

$$logit(\alpha) = \alpha_0 + \alpha_1 BC.RW.population.continuous + \alpha_2 study.RE \quad (26)$$

$$log(\gamma) = \gamma_0 + \gamma_1 BC.RW.population.continuous \quad (27)$$

B. Model priors

We allow each group to maintain a distinct random walk that varies from the population mean depending on the level of information they provide (sample sizes). Specifically, we assume a normal prior on the random walk for the alpha and gamma slopes as follows:

$$\alpha^1_{\mu,t} \sim N(\alpha^1_{\mu,t-1}, \sigma_{\alpha^1}{}^2) \quad (28)$$

$$\gamma^1_{\mu,t} \sim N(\gamma^1_{\mu,t-1}, \sigma_{\gamma^1}{}^2) \quad (29)$$

With weakly informative hyperpriors:

$$\alpha^1_{\mu,t=1} \sim N(0, \sigma = 0.2) \quad (30)$$

$$\gamma^1_{\mu,t=1} \sim N(0, \sigma = 0.2) \quad (31)$$

$$\sigma_{\alpha^1}{}^2 \sim IG(2, 0.5) \quad (32)$$

$$\sigma_{\gamma^1}{}^2 \sim IG(2, 0.5) \quad (33)$$

We assume similar priors for the alpha and gamma intercepts as for the corresponding fixed effects in the continuous and binary cohort base models. Specifically, we specified the following priors:

$$\alpha^0_{k,\mu} \sim t_{(1)}(\alpha^0_{k,\mu}, \sigma_{\alpha^0}{}^2) \quad (34)$$

$$\gamma^0_{k,\mu} \sim t_{(3)}(\gamma^0_{k,\mu}, \sigma_{\gamma^0}{}^2) \quad (35)$$

$$\alpha^0_{k,\mu} \sim Cauchy(0, 1) \quad (36)$$

$$\gamma^0_{k,\mu} \sim t_{(3)}(-3.9, 1) \quad (37)$$

$$\sigma_{\alpha^0}{}^2 \sim IG(2, 1) \quad (38)$$

$$\sigma_{\gamma^0}{}^2 \sim IG(2, 1) \quad (39)$$

Same priors for θ and study-level random effects (12 and 14&15)

3. RW model with group-specific trajectories and intercepts

A. Brief description

For this extension, we allow for race/ethnic specific differences in trajectories and baseline values for the random walks, as follows:

$$logit(\alpha) = \alpha_0 + \alpha_1 BC.RW.population + \alpha_2 BC.RW.group.deviation + \alpha_3 study.RE \quad (40)$$

$$\log(\gamma) = \gamma_0 + \gamma_1 BC.RW.population + \gamma_2 BC.RW.group.deviation \quad (41)$$

where $BC.RW.group.deviation$ denotes race/ethnic group-specific deviations from population trajectories

B. Model priors

Same priors for θ and study-level random effects (12 and 14&15) and all the same priors specified for model 2 (28-39), with additional weakly informative priors for the group-specific trajectory deviations:

$$\alpha^2_{k,\mu,t} \sim N(\alpha^2_{k,\mu,t-1}, \sigma^2_{\alpha^2}) \quad (42)$$

$$\gamma^2_{k,\mu,t} \sim N(\gamma^2_{k,\mu,t-1}, \sigma^2_{\gamma^2}) \quad (43)$$

with hyperpriors:

$$\alpha^2_{\mu,t=1} \sim N(0, \sigma_{\alpha^2}) \quad (44)$$

$$\gamma^2_{\mu,t=1} \sim N(0, \sigma_{\gamma^2}) \quad (45)$$

$$\sigma^2_{\alpha^2} \sim IG(2, 1) \quad (46)$$

$$\sigma^2_{\gamma^2} \sim IG(2, 1) \quad (47)$$

### C. Computing ages at half-maximum seroprevalence

Given equations 1-3, the distribution of age at infection conditional on ever being infected is Exponential(γ). Thus, the age at half-maximum seroprevalence, which is equivalent to the median age at acquisition in a closed population, can be expressed as $\frac{\ln(2)}{\gamma}$ under the simplest kappa=1 formulation (all models except binary cohort models 3-6). The age at half-maximum seroprevalence under flexible kappas (binary cohort models 3-6, see equations 16 & 19) can be expressed $\frac{(\ln(2))^{1/\kappa}}{\gamma}$

### D. Selecting optimal models

Among the binary cohort models, the three specifications with study-level random effects substantially outperformed those without according to their DIC and expected log predictive densities and were within about one standard error of one another (Supplementary Table S1). We therefore selected the most parsimonious of the three, the study random effects model with κ fixed at 1, as our main binary cohort model. Its expected log predictive density was 6.4 units lower than that of the best-performing model with group-specific κ and higher than that of the model with a common κ. Its estimated effective sample size was 7768 (95% CrI: 6327, 9410). The maximum PSRF across parameters was 1.01.

Among the continuous cohort models, we selected the model with population-wide (shared) random walk trajectories, group-specific intercepts and study random effects as it provided the best out-of-sample predictive capacity and demonstrated successful convergence (Supplementary Table S2). The maximum PSRF across parameters was 1.02, also well below the conventional threshold of 1.1. Further inspection of univariate and multivariate convergence statistics indicated satisfactory independent *and* joint parameter convergence (Supplementary Tables S4-S7). The estimated effective sample size was 10760 (95% CrI: 8349, 13707).

All trace and density plots are provided in Supplementary Figures S1, S2, S3-S6.

### E. Supplementary Tables and Figures

| model | DIC | Max PSRF | LOOCV ELPD | Difference in LOOCV ELPD from best model (SE)* |
|---|---|---|---|---|
| kappa_global_study_re | 1833.0 | 1.003 | -978.5 | −13.3 (9.41) |
| study_re | 1834.6 | 1.013 | -977.7 | −12.4 (9.45) |
| kappa_partial_study_re | 1839.9 | 1.052 | -971.3 | -6.00 (9.02) |
| base | 1842.1 | 1.015 | -1031.6 | −66.4 (16.0) |
| kappa_partial | 1842.9 | 1.179 | -1032.9 | −67.6 (16.0) |
| kappa_global | 1843.9 | 1.034 | -1032.6 | −67.3 (15.9) |

Supplementary Table S1: Model comparison under the binary cohort year analysis by DIC, maximum potential scale reduction factor (PSRF) and leave-one-out cross-validation (LOOCV) expected log predictive density. *compared to the best performing model, among all binary and continuous cohort models, with the highest LOOCV expected log predictive density. ELPD: expected log predictive density

| model | DIC | Max PSRF | LOOCV ELPD | Difference in LOOCV ELPD from best model (SE)* |
|---|---|---|---|---|
| RW_expanded_continuous_bc | 1827.1 | 1.039 | -978.9 | −13.6 (6.06) |
| RW_continuous_bc | 1843.4 | 1.017 | -965.3 | 0 |
| base_continuous_bc | 1861.6 | 4.422 | -1248.7 | −283 (66.6) |

Supplementary Table S2: Model comparison under the continuous cohort year analysis by DIC, maximum potential scale reduction factor (PSRF) and leave-one-out cross-validation (LOOCV) expected log predictive density. *compared to the model, among all binary and continuous cohort models, with the highest LOOCV expected log predictive density. ELPD: expected log predictive density

| Model | Group | Mean kappa | Median kappa | Lower 95% CrI | Upper 95% CrI | probability kappa > 1 |
|---|---|---|---|---|---|---|
| Global kappa model | Population-wide | 0.938 | 0.930 | 0.727 | 1.19 | 0.279 |
| Global kappa model with study RE | Population-wide | 1.05 | 1.05 | 0.880 | 1.26 | 0.701 |
| Group-specific kappa model | Hispanic | 1.03 | 1.02 | 0.759 | 1.35 | 0.544 |
| Group-specific kappa model | AI/AN | 0.796 | 0.690 | 0.300 | 1.78 | 0.267 |
| Group-specific kappa model | API | 1.29 | 1.18 | 0.409 | 2.86 | 0.644 |
| Group-specific kappa model | NHB | 0.669 | 0.654 | 0.427 | 1.00 | 0.0251 |
| Group-specific kappa model | NHW | 0.854 | 0.840 | 0.522 | 1.27 | 0.199 |
| Group-specific kappa model with study RE | Hispanic | 1.08 | 1.08 | 0.902 | 1.28 | 0.788 |
| Group-specific kappa model with study RE | AI/AN | 0.857 | 0.805 | 0.391 | 1.63 | 0.279 |
| Group-specific kappa model with study RE | API | 1.83 | 1.61 | 0.552 | 4.46 | 0.832 |
| Group-specific kappa model with study RE | NHB | 0.810 | 0.789 | 0.550 | 1.21 | 0.112 |
| Group-specific kappa model with study RE | NHW | 2.09 | 2.11 | 1.04 | 3.08 | 0.981 |

Supplementary Table S3: Kappa summary statistics for kappa-extended binary cohort models

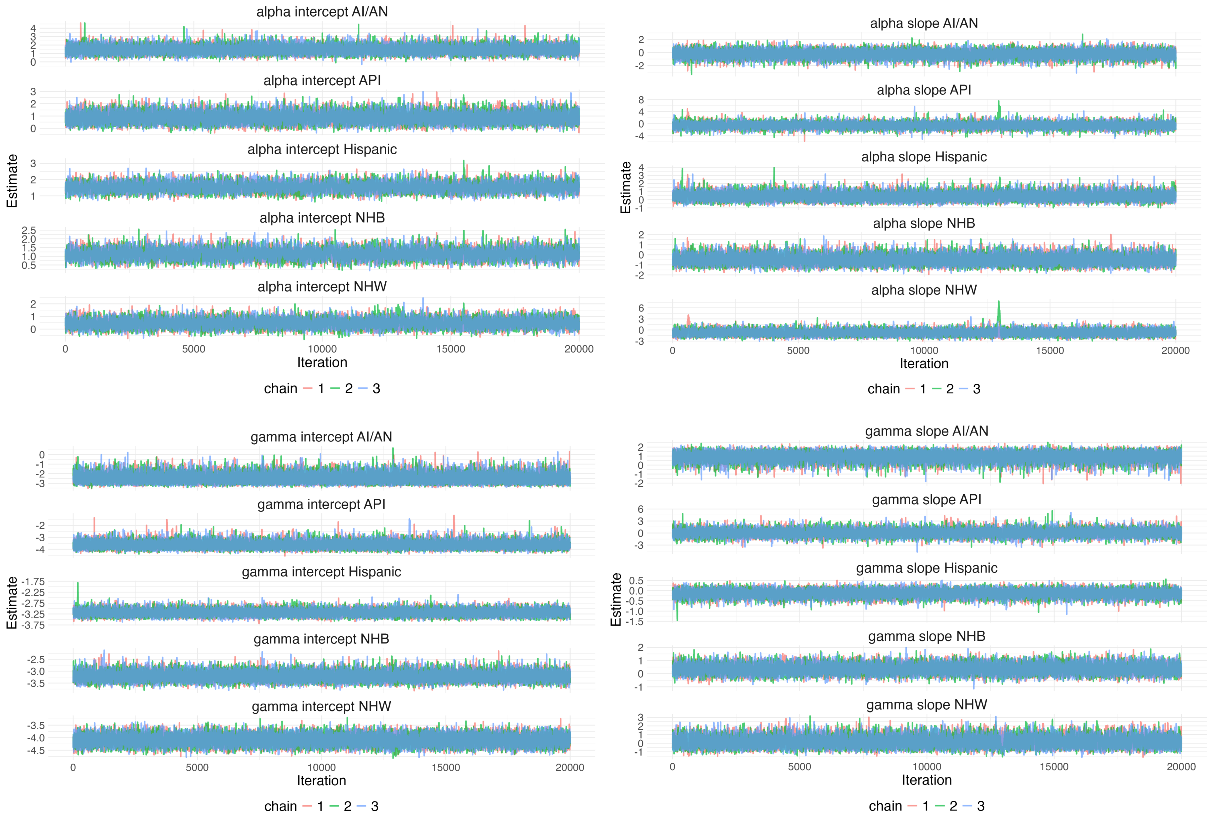


Supplementary Figure S1: Trace plots for key model parameters (parameter labels above each subplot) for the binary birth cohort main analysis. AI/AN: American Indian/Alaska Native, API: Asian and Pacific Islander, NHB: Non-Hispanic Black, NHW: Non-Hispanic White.

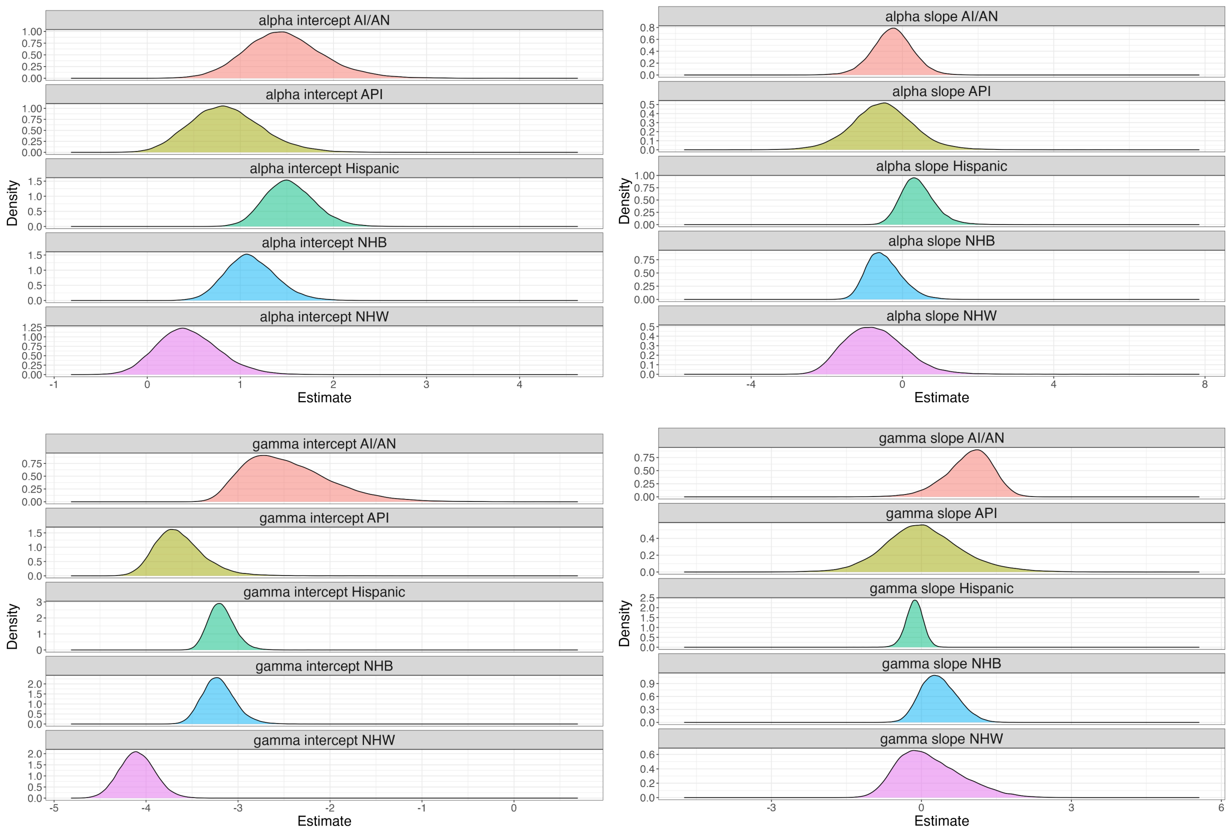


Supplementary Figure S2: Density plots for all model parameters (parameter labels above each subplot) for the binary birth cohort main analysis. AI/AN: American Indian/Alaska Native, API: Asian and Pacific Islander, NHB: Non-Hispanic Black, NHW: Non-Hispanic White.

| parameter | PSRF |
|---|---|
| log_alpha0_H | 1.00 |
| log_alpha0_AIAN | 1.00 |
| log_alpha0_API | 1.00 |
| log_alpha0_NHB | 1.00 |
| log_alpha0_NHW | 1.00 |
| log_alpha1_H | 1.00 |
| log_alpha1_AIAN | 1.00 |
| log_alpha1_API | 1.00 |
| log_alpha1_NHB | 1.00 |
| log_alpha1_NHW | 1.00 |
| log_gamma0_H | 1.00 |
| log_gamma0_AIAN | 1.00 |
| log_gamma0_API | 1.00 |
| log_gamma0_NHB | 1.00 |
| log_gamma0_NHW | 1.00 |
| log_gamma1_H | 1.00 |
| log_gamma1_AIAN | 1.00 |
| log_gamma1_API | 1.00 |
| log_gamma1_NHB | 1.00 |
| log_gamma1_NHW | 1.00 |

Supplementary Table S4: Univariate convergence statistics - potential scale reduction factors (PSRF) by parameter for the binary birth cohort main analysis. AI/AN: American Indian/Alaska Native, API: Asian and Pacific Islander, NHB: Non-Hispanic Black, NHW: Non-Hispanic White.

| **parameter** | **PSRF** |
|---|---|
| mu_alpha0 | 1.00 |
| mu_alpha1 | 1.00 |
| mu_gamma0 | 1.00 |
| mu_gamma1 | 1.00 |
| var_alpha0 | 1.00 |
| var_alpha1 | 1.01 |
| var_gamma0 | 1.00 |
| var_gamma1 | 1.00 |
| covar_alpha0_1 | 1.00 |
| covar_alpha0_gamma0 | 1.00 |
| covar_gamma0_alpha1 | 1.00 |
| covar_gamma1_alpha0 | 1.00 |
| covar_gamma1_alpha1 | 1.00 |
| covar_gamma0_1 | 1.00 |
| overdispersion_parameter | 1.00 |
| sd_study | 1.00 |

Supplementary Table S5: Multivariate convergence statistics - potential scale reduction factors (PSRF) by parameter for the binary birth cohort main analysis.

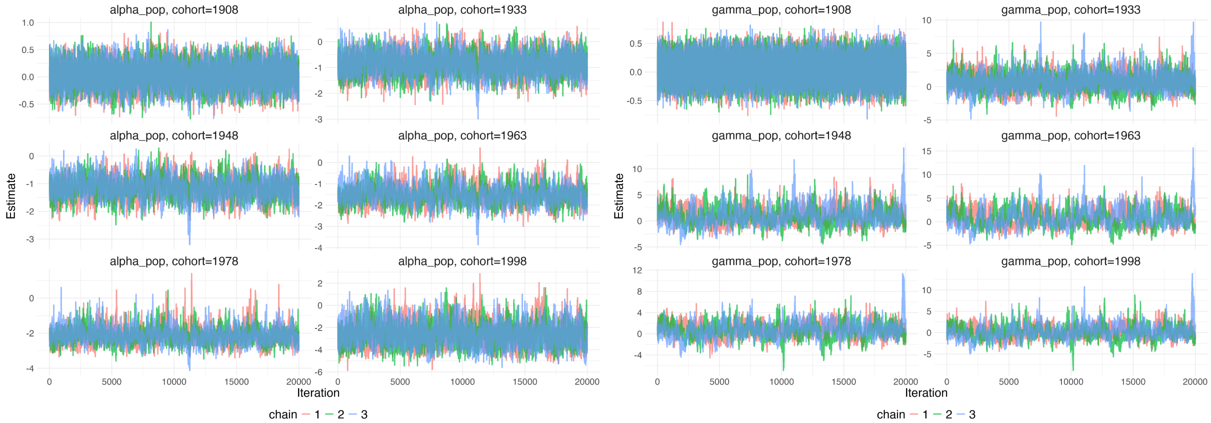


Supplementary Figure S3: Trace plots for key population-wide model parameters (parameter labels above each subplot) for the individual birth cohort year main analysis.

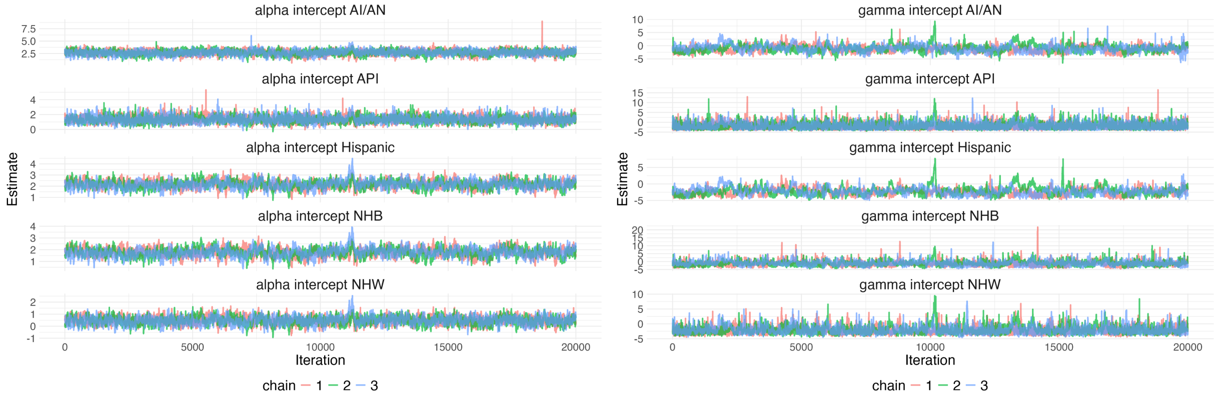


Supplementary Figure S4: Trace plots for race/ethnic group-specific model parameters (parameter labels above each subplot; each row corresponds to the Hispanic, AI/AN, API, NHB, and NHW populations, alpha (left) and gamma (right) trajectory deviations) for the individual birth cohort year main analysis. AI/AN: American Indian/Alaska Native, API: Asian and Pacific Islander, NHB: Non-Hispanic Black, NHW: Non-Hispanic White.

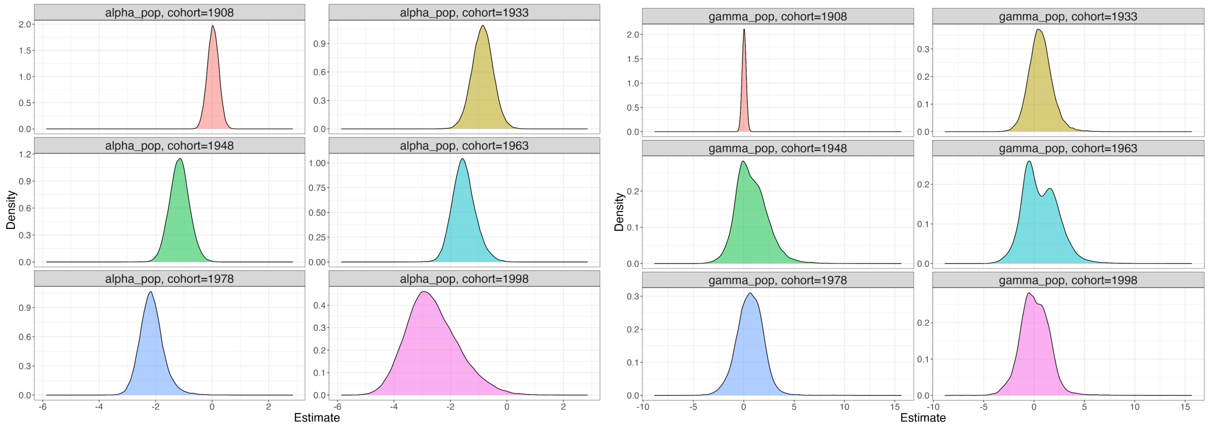


Supplementary Figure S5: Density plots for key population-wide model parameters (parameter labels above each subplot) for the individual birth cohort year main analysis.

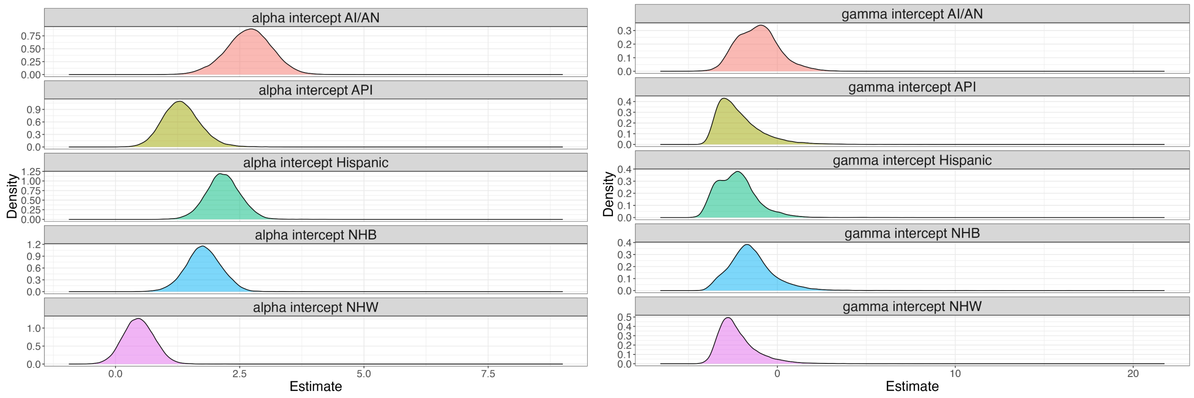


Supplementary Figure S6: Density plots for race/ethnic group-specific model parameters (parameter labels above each subplot; each row corresponds to the Hispanic, AI/AN, API, NHB, and NHW populations, alpha (left) and gamma (right) trajectory deviations) for the individual birth cohort year main analysis. AI/AN: American Indian/Alaska Native, API: Asian and Pacific Islander, NHB: Non-Hispanic Black, NHW: Non-Hispanic White.

| parameter | PSRF |
|---|---|
| log_alpha0_H | 1.00 |
| log_alpha0_AIAN | 1.00 |
| log_alpha0_API | 1.00 |
| log_alpha0_NHB | 1.00 |
| log_alpha0_NHW | 1.00 |
| log_gamma0_H | 1.02 |
| log_gamma0_AIAN | 1.01 |
| log_gamma0_API | 1.01 |
| log_gamma0_NHB | 1.01 |
| log_gamma0_NHW | 1.01 |

Supplementary Table S6: Univariate convergence statistics - potential scale reduction factors (PSRF) by parameter for the individual birth cohort year main analysis. AI/AN: American Indian/Alaska Native, API: Asian and Pacific Islander, NHB: Non-Hispanic Black, NHW: Non-Hispanic White.

| parameter | PSRF |
|---|---|
| mu_alpha0 | 1.00 |
| mu_gamma0 | 1.02 |
| overdispersion_parameter | 1.00 |
| sd_rw_alpha_pop | 1.00 |
| sd_rw_gamma_pop | 1.01 |
| sd_study | 1.00 |

Supplementary Table S7: Multivariate convergence statistics - potential scale reduction factors (PSRF) by parameter for the individual birth cohort year main analysis.

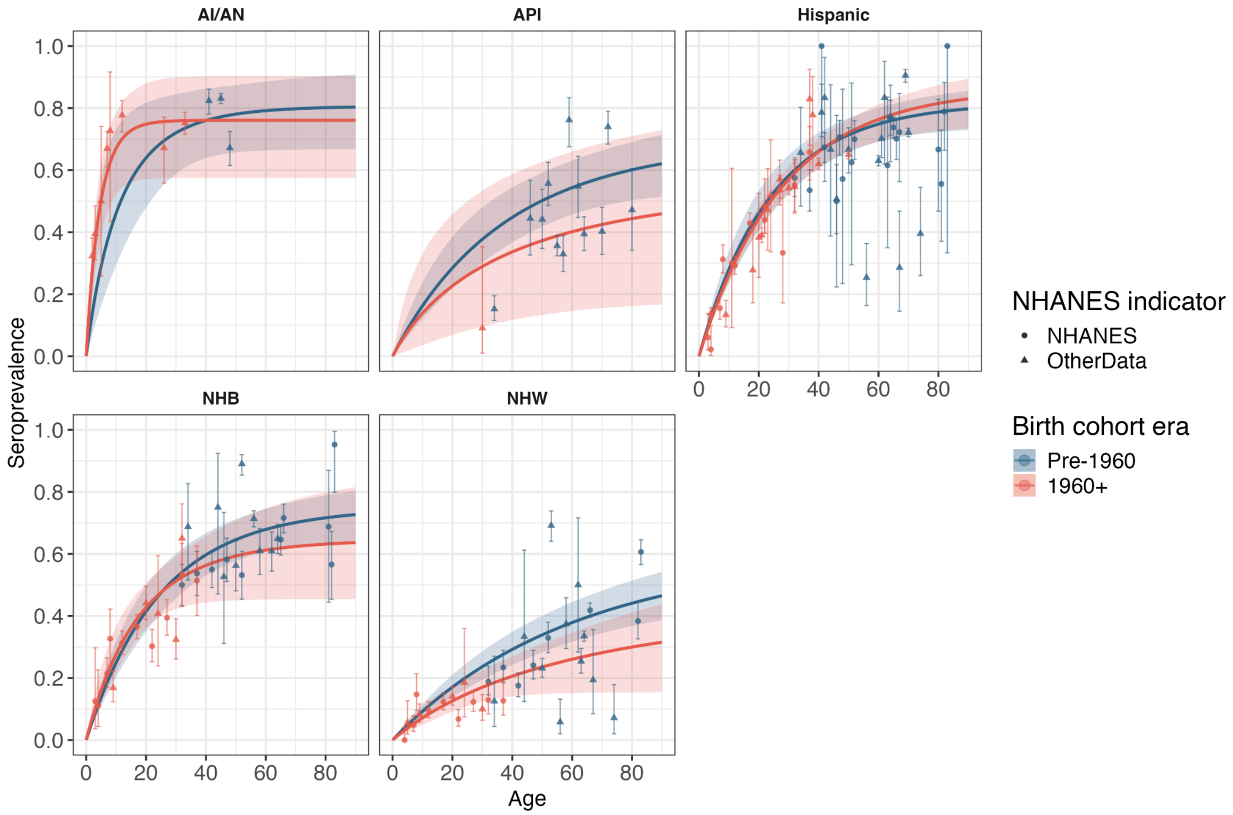


Supplementary Figure S7: Model-predicted and observed prevalence by age, birth cohort era, and race/ethnic group (birth cohort binary designation of pre-1960 and 1960+) under the selected binary cohort model (common kappa with study random effects). Curves indicate posterior means and shaded regions indicate 95% credible intervals. Points and error bars indicate observed means and 95% confidence intervals. Circles denote NHANES data and triangles denote data from the other studies captured in the scoping review. AI/AN: American Indian/Alaska Native, API: Asian and Pacific Islander, NHB: Non-Hispanic Black, NHW: Non-Hispanic White.

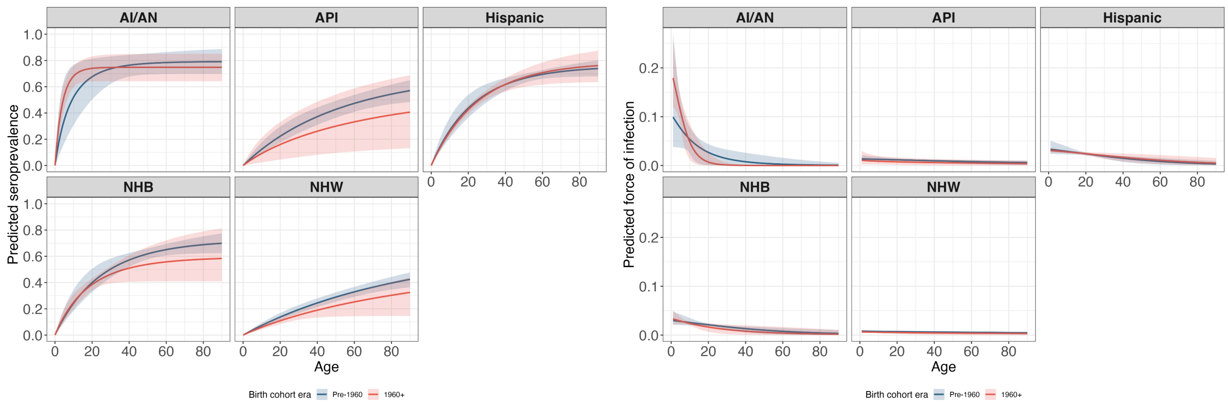


Supplementary Figure S8: Model-predicted prevalence (left) and forces of infection (right) by age, birth cohort year and race/ethnic group under the base model. AI/AN: American Indian/Alaska Native, API: Asian and Pacific Islander, NHB: Non-Hispanic Black, NHW: Non-Hispanic White.

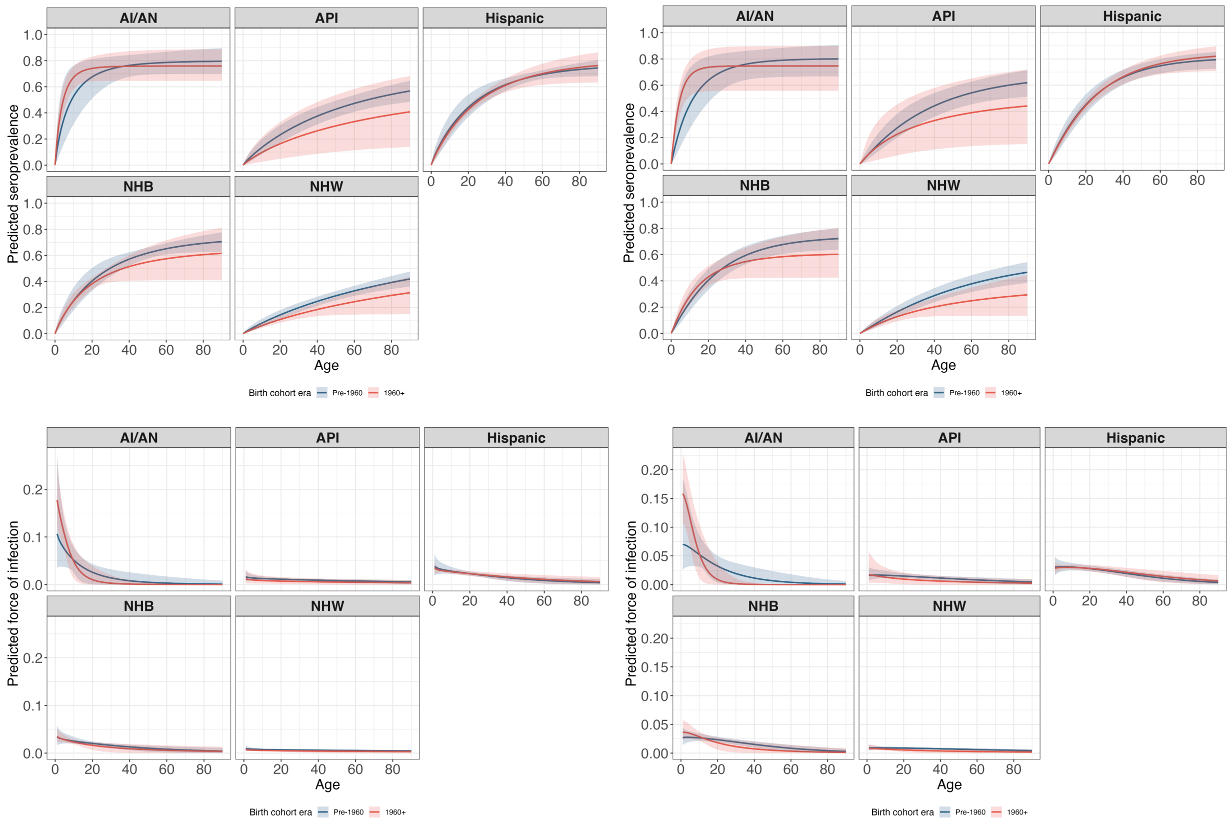


Supplementary Figure S9: Model-predicted prevalence by age, birth cohort year and race/ethnic group under the alternative binary cohort models: common kappa model (top left) and common kappa model with study random effects (top right). Model-predicted forces of infection by age, birth cohort year, and race/ethnic group under the alternative binary cohort models: common kappa model (bottom left) and common kappa model with study random effects (bottom right). AI/AN: American Indian/Alaska Native, API: Asian and Pacific Islander, NHB: Non-Hispanic Black, NHW: Non-Hispanic White.

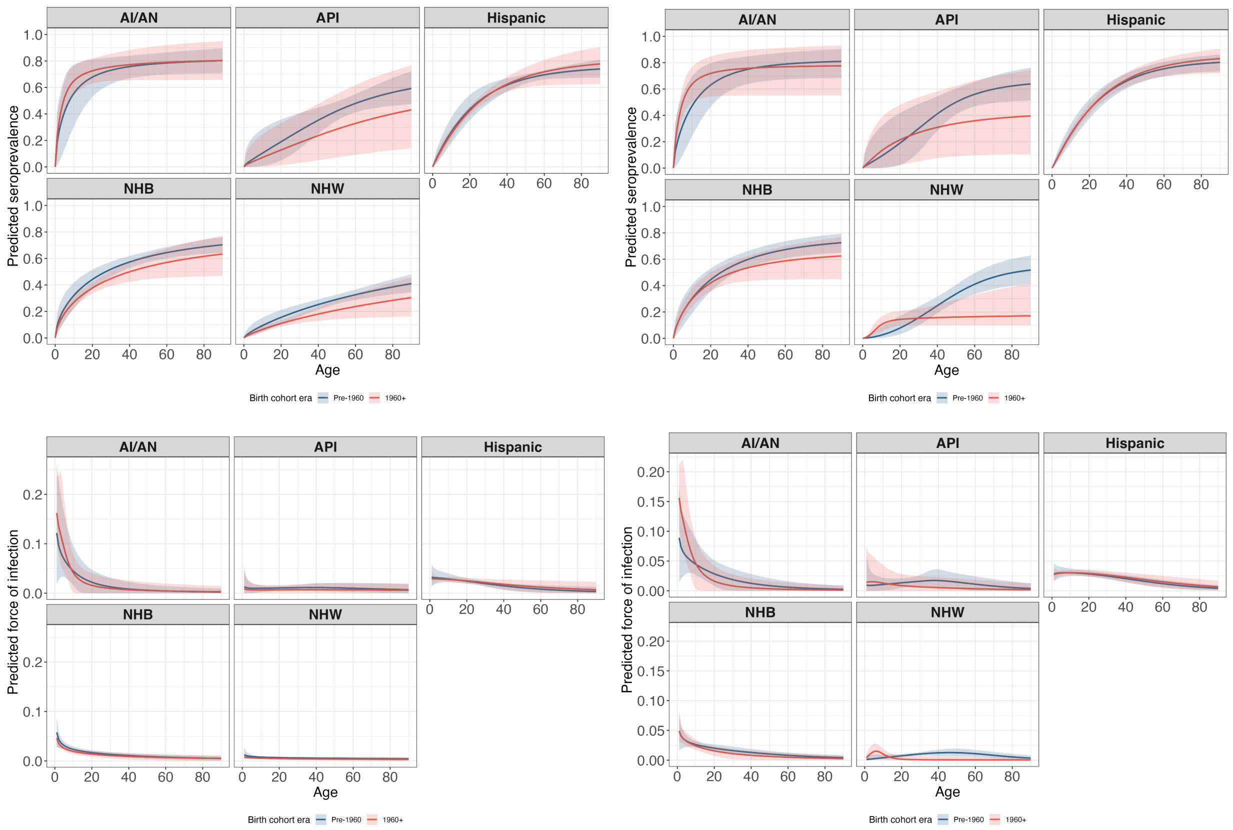


Supplementary Figure S10: Model-predicted prevalence by age, birth cohort year and race/ethnic group under the alternative binary cohort models: group-specific kappa model (top left) and group-specific kappa model with study random effects (top right). Model-predicted forces of infection by age, birth cohort year and race/ethnic group under the alternative binary cohort models: group-specific kappa model (bottom left) and group-specific kappa model with study random effects (bottom right). AI/AN: American Indian/Alaska Native, API: Asian and Pacific Islander, NHB: Non-Hispanic Black, NHW: Non-Hispanic White.

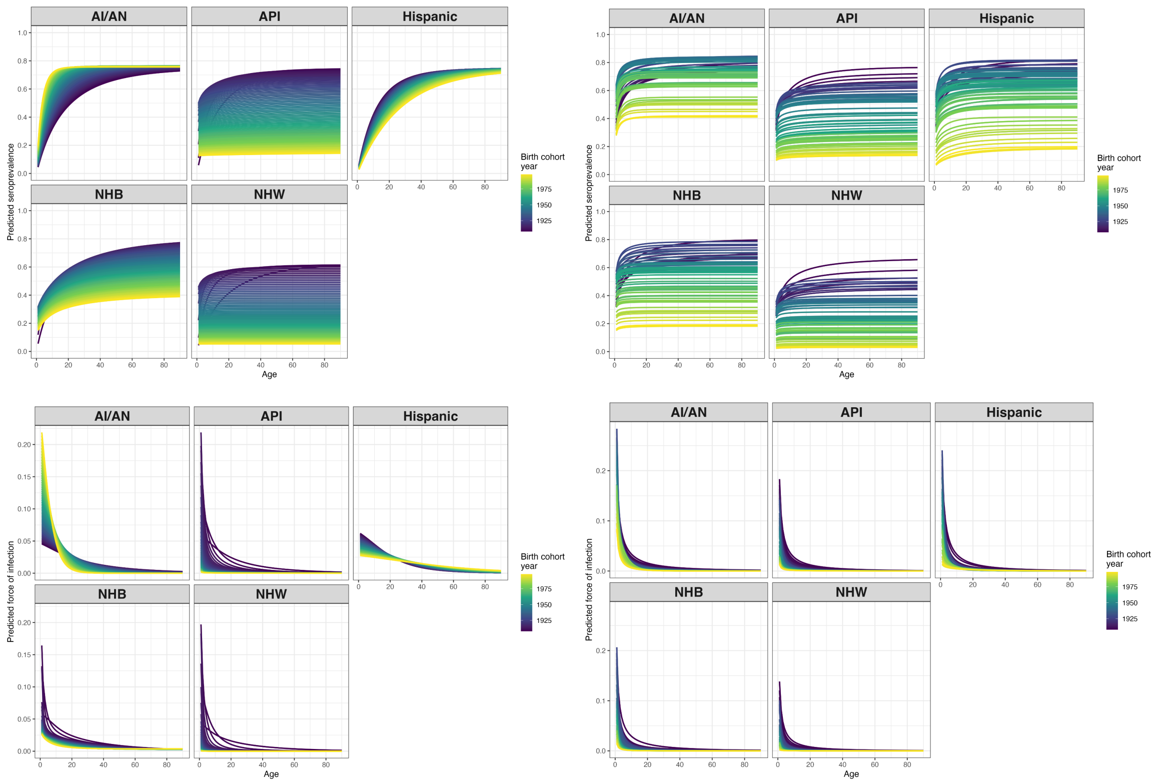


Supplementary Figure S11: Model-predicted prevalence by age, birth cohort year and race/ethnic group under the alternative continuous birth cohort models: base model (top left) and expanded RW model with race-specific intercepts and trajectories (top right). Model-predicted forces of infection by age, birth cohort year and race/ethnic group under the alternative (non-selected) continuous birth cohort models: base model (bottom left) and expanded RW model with race-specific intercepts and trajectories (bottom right). Curves indicate posterior means. AI/AN: American Indian/Alaska Native, API: Asian and Pacific Islander, NHB: Non-Hispanic Black, NHW: Non-Hispanic White.
